# Upregulation of Human Endogenous Retroviruses in Bronchoalveolar Lavage Fluid of COVID-19 Patients

**DOI:** 10.1101/2020.05.10.20096958

**Authors:** Konstantina Kitsou, Anastasia Kotanidou, Dimitrios Paraskevis, Timokratis Karamitros, Aris Katzourakis, Richard Tedder, Tara Hurst, Spyros Sapounas, Athanassios Kotsinas, Vassilis Gorgoulis, Vana Spoulou, Sotirios Tsiodras, Pagona Lagiou, Gkikas Magiorkinis

## Abstract

**Background:** Severe COVID-19 pneumonia has been associated with the development of intense inflammatory responses during the course of infections with SARS-CoV-2. Given that Human Endogenous Retroviruses (HERVs) are known to be activated during and participate in inflammatory processes, we examined whether HERV dysregulation signatures are present in COVID-19 patients.

**Results:** By comparing transcriptomes of Peripheral Blood Monocytes (PBMCs) and Bronchoalveolar Lavage Fluid (BALF) from patients and normal controls we have shown that HERVs are intensely dysregulated in BALF, but not in PBMCs. In particular, upregulation in the expression of multiple HERV families was detected in BALF samples of COVID-19 patients, with HERV-W being the most highly upregulated family among the families analysed. In addition, we compared the expression of HERVs in Human Bronchial Epithelial Cells (HBECs) without and after senescence induction in an oncogene-induced senescence model, in order to quantitatively measure changes in the expression of HERVs in bronchial cells during the processes of cellular senescence.

**Conclusions:** This apparent difference of HERV dysregulation between PBMCs and BALF warrants further studies in involvement of HERVs in inflammatory pathogenetic mechanisms as well as exploration of HERVs as potential biomarkers for disease progression. Furthermore, the increase in the expression of HERVs in senescent HBECs in comparison to non-induced HBECs provides a potential link for increased COVID-19 severity and mortality in aged populations.

## Background

Human Endogenous Retroviruses (HERVs) are the remnants of ancient retroviral infections that infiltrated the germlines of our deep in time ancestors and integrated within their genomes. Through Mendelian inheritance and evolutionary processes spanning millions of years they now occupy around 8% of the human genome (1). Their evolutionary history based on analyses of molecular sequences suggests that at least 30 independent colonization events generated an equivalent number of closely related retroviral integrations known as families (2).

The vast majority of HERV integrations have accumulated mutations that effectively incapacitated their proliferative functionality, however the these integrants can still be transcribed and encode proteins (3). HERV expression is usually silenced through numerous post-transcriptional mechanisms, but can be upregulated in certain diseases and conditions such as cancer and inflammation (4,5). The retroviral involvement in inflammatory processes has been shown to occur through over-production of nucleic acids and proteins that interfere with a variety of innate immune response and inflammatory pathways (6).

COVID-19 emerged in late 2019 in Wuhan City and from China spread around the globe generating the most intensive pandemic responses within the last 50 years leading to significant socioeconomic disruption. The disease is caused by a zoonotic coronavirus, now known as SARS-CoV-2, which in a proportion of individuals will cause severe pneumonia, respiratory and multi-organ failure referred to as Corona virus infectious disease 2019 (COVID-19) (7). For severe COVID-19 it has been suggested that a critical component of the pathophysiology is a severe inflammatory response driven by cytokine release (8).

Here we aimed to explore whether HERV expression is upregulated in patients with COVID-19 as this could be consistent with an involvement of HERVs in inflammatory responses such as the production of interferonogenic nucleic acids and inflammatory proteins. In the initial study, in which the COVID-19 patients’ RNAseq data were produced, the authors correlated the increased expression of proinflammatory cytokines and cytokine receptors in Bronchoalveolar Lavage Fluid (BALF) samples of COVID-19 patients when compared to healthy subjects, and related this finding to the cytokine storm and disease severity in these patients (9). In addition, we aimed to determine the differences between the expression of HERVs after oncogene-induced senescence of Human Bronchial Epithelial Cells (HBECs) in a non-malignant senescence model and the expression of HERVs in non-induced HBECs (10,11). In this way, we aim to determine the effect of senescence on the expression of HERVs as a potential component of the increased inflammatory burden of senescence in bronchial cells (10). We hypothesized that the expression of HERVs is augmented in the state of senescence and this could offer a plausible explanation for the increased COVID-19 severity and mortality observed as the patient age increases.

We find that multiple HERV families are upregulated in BALF, but not in Peripheral Blood Monocytes (PBMCs), in patients with COVID-19 compared to healthy individuals. The findings merit further study regarding the potential involvement of HERVs in COVID-19.

## Results

### Expression of HERVs in BALF samples and PBMCs from COVID-19 patients in comparison to healthy controls

Transcription of HERVs was normalized by means of two housekeeping genes, succinate dehydrogenase (SDHA) and hypoxanthine phosphoribosyl transferase 1 (HPRT1). Upregulation in the expression of endogenous retroviral elements was observed in the BALF samples of COVID-19 patients in comparison to healthy controls. In particular, in the normalized expression, with the SDHA expression, statistically significant upregulation in the BALF samples of COVID-19 patients in comparison to healthy controls was found in the expression of HERV-K (HML-1) (2.94 fold change, p=0.047), HERV-K (HML-2) (2.59 fold change, p=0.041), HERV-K (HML-3) (4.95 fold change, p<0.001), HERV-K (HML-5) (4.5 fold change, p=0.002), HERV-K (HML-6) (2 fold change, p=0.005), HERV-W (8.33 fold change, p<0.001), HERV-H (6.64 fold change, p<0.001), ERV-L (6.33 fold change, p<0.001), HERV-9 (5.44 fold change, p=0.004), HERV-E (4.65 fold change, p<0.001), HERV-FRD (4.35 fold change, p=0.009) (Figure 1 and Figure 2). No statistically significant differences were observed in the expression of HERV-K (HML-4) (p=0.709) and HERV-I (p=0.053).

**Figure 1:**
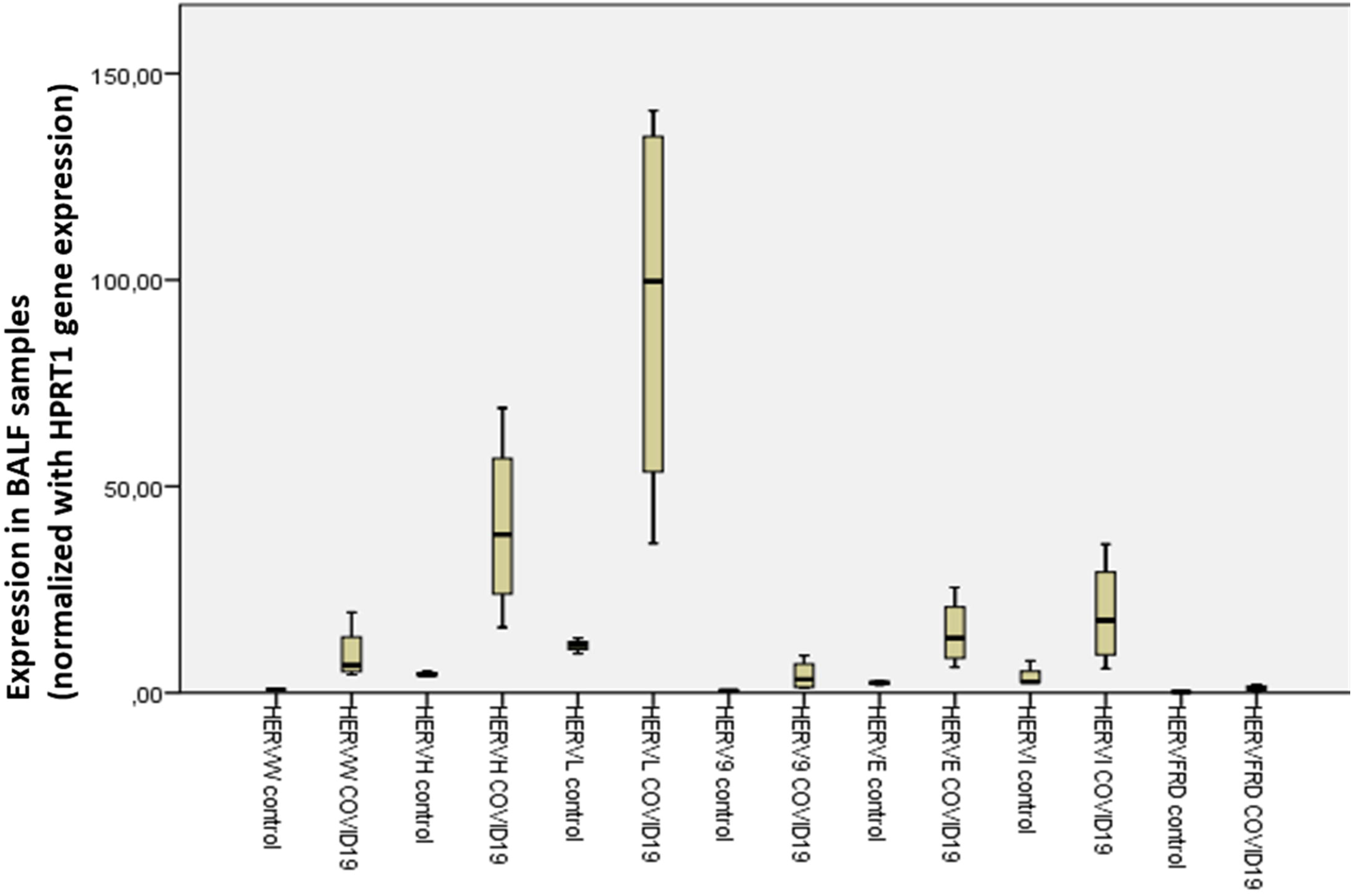
Upregulation of HML-1, HML-2, HML-3, HML-5, HML-6 in BALF of COVID-19 patients (SDHA normalization) Legend:: Expression of HERV-K (HML-1), HERV-K (HML-2), HERV-K (HML-3), HERV-K (HML-4), HERV-K (HML-5), HERV-K (HML-6) in BALF of healthy controls and COVID-19 patients normalized with the expression of SDHA gene. Statistically significant upregulation in the BALF samples of COVID-19 patients in comparison to healthy controls was observed in the expression of HML-1 (2.94 fold change, p=0.047), HML-2 (2.59 fold change, p=0.041), HML-3 (4.95 fold change, p<0.001), HML-5 (4.5 fold change, p=0.002), HML-6 (2 fold change, p=0.005).

**Figure 2:**
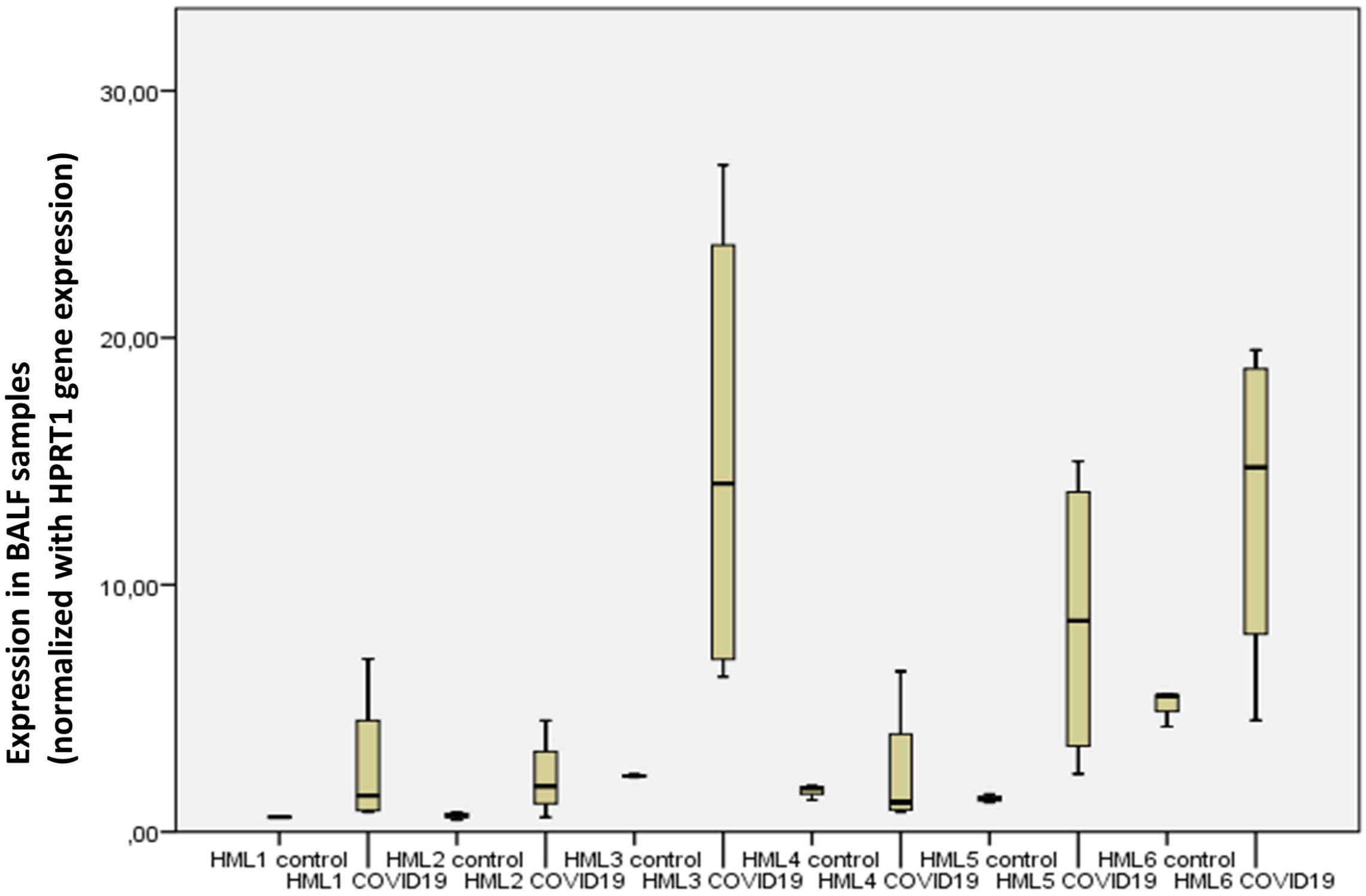
Upregulation of HERV-W, HERV-H, ERV-L, HERV-9, HERV-E, HERV-FRD the BALF of COVID-19 patients (SDHA normalization) Legend: Expression of HERV-W, HERV-H, ERV-L, HERV-9, HERV-E, HERV-I and HERV-FRD in BALF of healthy controls and COVID-19 patients normalized with the expression of SDHA gene. Statistically significant upregulation in the BALF samples of COVID-19 patients in comparison to healthy controls was observed in the expression of HERV-W (8.33 fold change, p<0.001), HERV-H (6.64 fold change, p<0.001), ERV-L (6.33 fold change, p<0.001), HERV-9 (5.44 fold change, p=0.004), HERV-E (4.65 fold change, p<0.001), HERV-FRD (4.35 fold change, p=0.009).

In the normalization with the HPRT1 expression, the upregulation in the expression of HERVs in COVID-19 patients was statistically significant in the expression of HERV-K (HML-3) (6.79 fold change, p=0.017), HERV-K (HML-5) (6.41 fold change, p=0.033), HERV-W (11.06 fold change, p=0.005), HERV-H (8.73 fold change, p=0.006), ERV-L (8.25 fold change, p=0.006), HERV-9 (8.18 fold change, p=0.036), HERV-E (6.05 fold change, p=0.007), HERV-FRD (5.31 fold change, p=0.014) and HERV-I (4.47 fold change, p=0.014), while no statistically significant upregulation was observed in the expression of HERV-K (HML-1), HERV-K (HML-2), HERV-K (HML-4), and HERV-K (HML-6) (Figure 3 and Figure 4).

**Figure 3:**
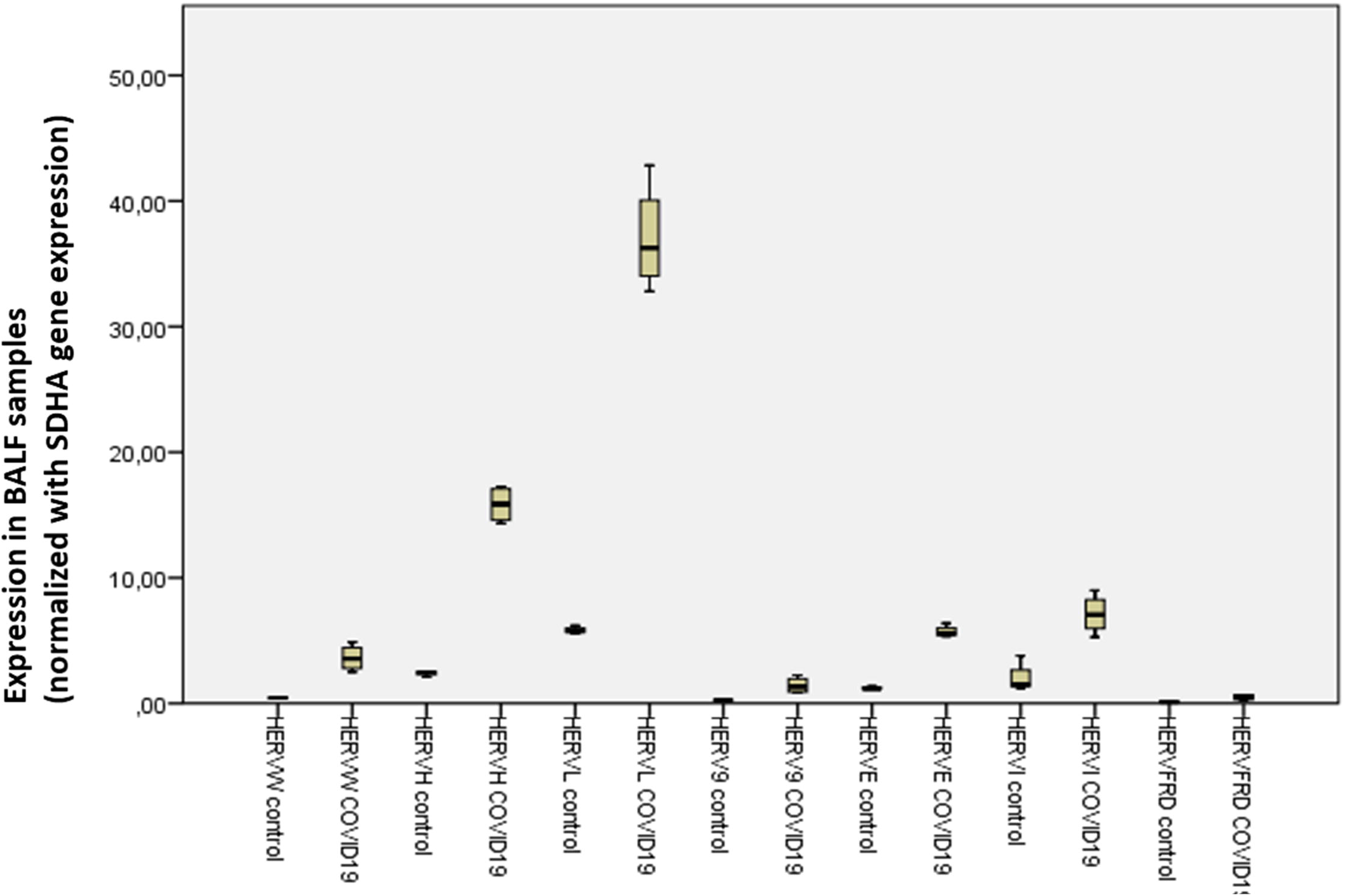
Upregulation of HML-3 and HML-5 in the BALF of COVID-19 patients (HPRT1 normalization) Legend: Expression of HERV-K (HML-1), HERV-K (HML-2), HERV-K (HML-3), HERV-K (HML-4), HERV-K (HML-5), HERV-K (HML-6) in BALF of healthy controls and COVID-19 patients normalized with the expression of HPRT1 gene. Statistically significant upregulation in the BALF samples of COVID-19 patients in comparison to healthy controls was observed in the expression of HML-3 (6.79 fold change, p=0.017), HML-5 (6.41 fold change, p=0.033).

**Figure 4:**
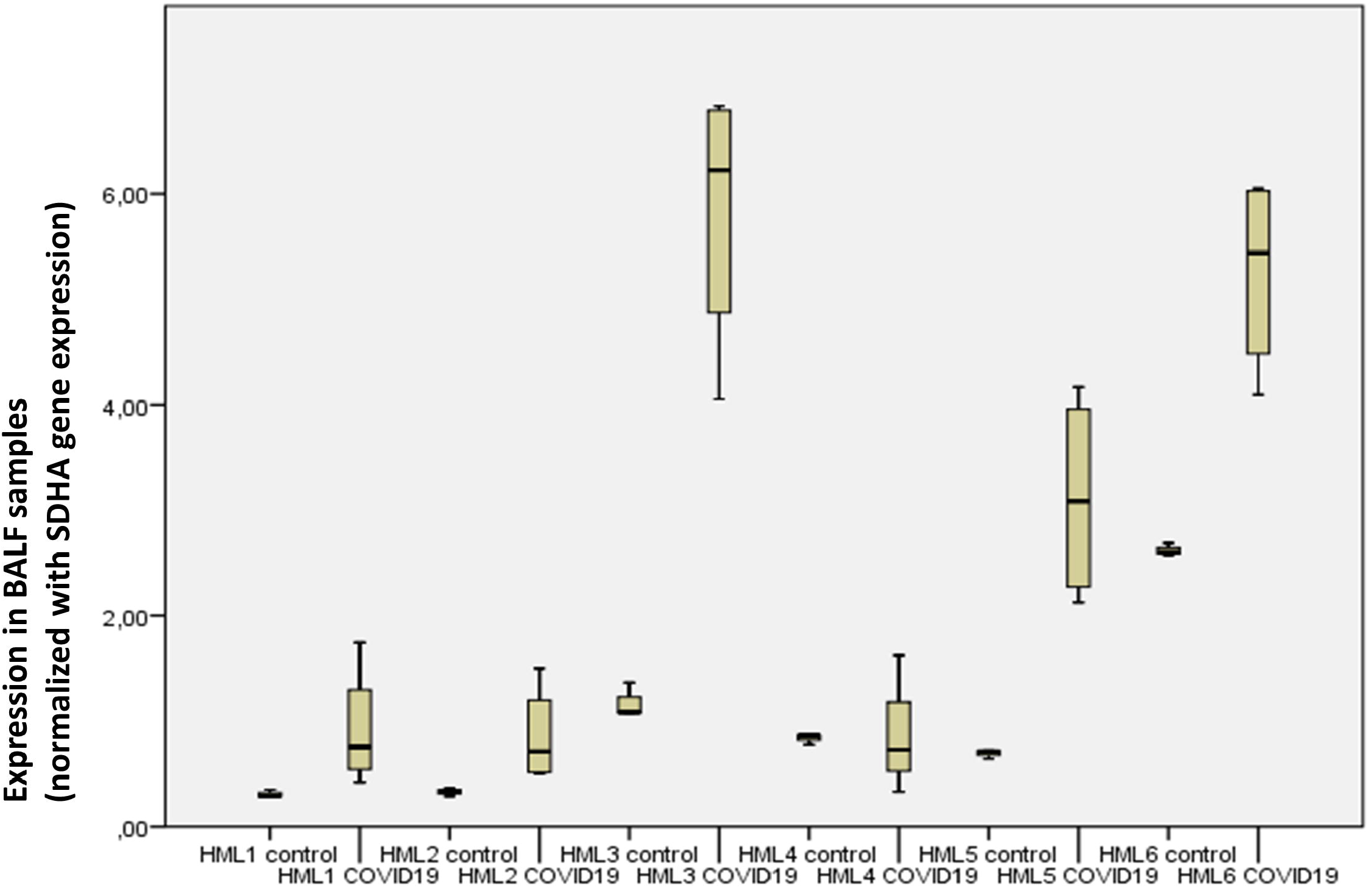
Upregulation of HERV-W, HERV-H, ERV-L, HERV-9, HERV-E, HERV-FRD and HERV-I in BALF of COVID-19 patients (HPRT1 normalization) Legend: Expression of HERV-W, HERV-H, HERV-L, HERV-9, HERV-E, HERV-I and HERV-FRD in BALF of healthy controls and COVID-19 patients normalized with the expression of HPRT1 gene. Statistically significant upregulation in the BALF samples of COVID-19 patients in comparison to healthy controls was observed in the expression of HERV-W (11.06 fold change, p=0.005), HERV-H (8.73 fold change, p=0.006), ERV-L (8.25 fold change, p=0.006), HERV-9 (8.18 fold change, p=0.036), HERV-E (6.05 fold change, p=0.007), HERV-FRD (5.31 fold change, p=0.014) and HERV-I (4.47 fold change, p=0.014)

In the normalization with the SDHA expression in the PBMCs datasets, statistically significant downregulation of the expression of ERV-L (0.85 fold change, p=0.029) and HERV-FRD (0.64 fold change, p=0.049) was observed in COVID-19 patients’ PBMCs in comparison to PBMCs of healthy donors. No statistically significant differences were observed in the expression of HERV-K (HML-1), HERV-K (HML-2), HERV-K (HML-3), HERV-K (HML-4), HERV-K (HML-5), HERV-K (HML-6), HERV-W, HERV-H, HERV-9, HERV-E, and HERV-I (Figure 5 and Figure 6 in Additional Files 1 and 2 respectively).

In the normalized expression with the HPRT1 expression, statistically significant downregulation was observed in the expression of HERV-K (HML-1) in the PBMCs of COVID-19 patients in comparison to PBMCs of healthy controls (0.74 fold change, p=0.007). No statistically significant differences were observed in the expression of HERV-K (HML-2), HERV-K (HML-3), HERV-K (HML-4), HERV-K (HML-5), HERV-K (HML-6), HERV-W, HERV-H, ERV-L, HERV-9, HERV-E, HERV-FRD and HERV-I (Figure 7 and Figure 8 in Additional Files 3 and 4 respectively).

### Expression of HERVs in non-induced and induced (senescent) HBECs

Transcription of HERVs was normalized by means of two housekeeping genes, succinate dehydrogenase (SDHA) and hypoxanthine phosphoribosyl transferase 1 (HPRT1). Upregulation in the expression of endogenous retroviral elements was recognised in the senescence induced HBECs in comparison to non-induced HBECs. After the normalization with the expression of SDHA gene, statistically significant upregulation in the induced HBECs was detected for HERV-K (HML-1) (1.91 fold change, p<0.001), HERV-K (HML-2) (2.29 fold change, p<0.001), HERV-K (HML-3) (1.36 fold change, p=0.001), HERV-K (HML-5) (1.67 fold change, p<0.001), HERV-K (HML-6) (1.3 fold change, p=0.001), HERV-W (1.9 fold change, p<0.001), ERV-L (1.86 fold change, p<0.001), HERV-9 (1.74 fold change, p<0.001), HERV-E (3.38 fold change, p<0.001). No statistically significant differences were observed in the expression of HERV-H, HERV-I and HERV-FRD.

After the normalization with the expression of HPRT1 gene, statistically significant upregulation in the induced HBECs was detected for HERV-K (HML-1) (1.57 fold change, p<0.001), HERV-K (HML-2) (1,98 fold change, p<0.001), HERV-K (HML-3) (1.16 fold change, p=0.047), HERV-K (HML-5) (1.43 fold change, p<0.001), HERV-W (1.61 fold change, p<0.001), ERV-L (1.57 fold change, p<0.001), HERV-9 (1.51 fold change, p<0.001), HERV-E (3.57 fold change, p<0.001). No statistically significant differences were observed in the expression of HERV-K (HML-6), HERV-H and HERV-FRD.

Downregulation in the expression of senescent HBECs in comparison to non-induced HBECs was recognised in the expression of HERV-K (HML-4) after the normalization with SDHA and HPRT1 gene (0.7 fold change, p=0.011 and 0.61 fold change, p<0.001 respectively) and in the expression of HERV-I after normalizing with the expression of HPRT1 gene (0.7 fold change, p<0.001).

## Discussion

SARS-CoV-2 infection is either mild or asymptomatic in the majority of the infections (7,12). Age and underlying comorbidities like pulmonary and/or cardiovascular disease are major risk factors for severe COVID-19 (13). However, the underlying causes of the severity of the infection are not fully understood. Here we have explored whether HERVs are dysregulated in patients with COVID-19 as this could provide evidence in favour of the hypothesis that they could be implicated in the severity of the disease. Based on transcriptomic data we have found that multiple HERV families are upregulated in BALF, but not in PBMCs of COVID-19 patients. Our findings combined with data showing that entry factors for SARS-CoV-2 are co-expressed with innate immunity genes in respiratory cells suggests that HERV upregulation in BALF might indeed be relevant to local rather than systematic immunity responses (14).

While HERVs are not commonly expressed throughout life, they have on the other hand been shown to have a dual role with respect to inflammation. They are upregulated by inflammatory pathways (15), but also ERV proteins and nucleic acids may trigger inflammatory responses (16). For example, we have previously shown that IFI-16 has a broad range of ssDNA targets, thus if HERVs or other transposable elements are upregulated and reverse transcribed as a result of COVID-19, this could amplify inflammatory responses (17). In our analyses HERV-W was the most highly upregulated family (up to 11x times higher). Interestingly, HERV-W *env* has been shown to be associated with proinflammatory transcriptional signatures and induce proinflammatory responses (18). Whether the upregulation of HERV-W RNA seen in transcriptomes results in production of HERV-W proteins remains to be determined.

Crucially, HERV expression seems to have an association with age, which is the strongest risk factor for severe COVID-19. Differential expression of HERVs has been described when comparing older with younger adults (19). Upregulation of endogenous retroviruses as a result of aging has been studied in mice whereas failure of epigenetic mechanisms seems to be the driving force for upregulation of endogenous retroviruses with age (20). Hence, it could be hypothesized that a possible lack of epigenetic regulation of HERVs in older individuals, may result in HERV expression overdrive during COVID-19 which in turn results in massive pro-inflammatory responses (21). This association between aging and increased expression of endogenous retroviral elements is confirmed by our findings. In the frameworks of this study an upregulation in the expression of most HERV families included in our analyses is demonstrated in senescence induced HBECs in comparison to non-induced. These results indicate that the increased expression of endogenous retroviral elements in senescent bronchial epithelial cells possibly induces local chronic inflammation, as a hallmark of immunosenescence and enhance the hypothesis of local rather than systematic inflammatory reactions. This hypothesis provides a plausible explanation for the increased COVID-19 severity and mortality as age increases.

Furthermore, recently, COVID-19 exposure has been linked to hyperinflammatory shock in peadiatric patients with manifestations that resemble Kawasaki Disease (both classic and incomplete), thus COVID-19 was linked to an unexpected increase of cases of the syndrome (22). Intriguingly, in a case series of Kawasaki peadiatric patients linked to the COVID-19 epidemic, HERV levels were detectable in the microbiology results of one of the eight afflicted children described in this case series (22). This finding could potentially indicate a further role of HERVs as an inflammatory trigger upon SARS-CoV-2 infection.

One limitation of our study is the small number of samples; thus, our findings need to be replicated in more patients, but also need to be followed up by functional studies. Nevertheless, our findings might also (at least partially) provide a working hypothesis to explain the mechanism of action of Lopinavir/ritonavir which has been shown to inhibit the expression of at least one HERV family (HK2) (23). Interestingly, there is also evidence of the potential benefit of antiretroviral treatment against SARS-CoV-2 infection, as there is an ongoing phase 2b/3 Clinical Trial for the assessment of the effectiveness of leronlimab, a CCR5 antagonist, against COVID-19 (24).

We find that multiple HERV families are upregulated in Bronchoalveolar Lavage Fluid (BALF), but not in PBMCs, in patients with COVID-19 compared to healthy individuals. Furthermore, we were able to identify upregulation in the expression of HERVs in senescence induced HBECs in comparison to non-induced cells, a fact that indicates the potential role of increased endogenous retroviral expression as a mediator of inflammatory reactions in older individuals that are at increased risk due to the disease. The findings merit further study regarding the potential involvement of HERVs in COVID-19. It thus seems feasible that should HERV expression be aetiologically linked to COVID-19 this expression could be a therapeutic target that to minimize the likelihood of severe Covid-19 and death. It remains to be seen if HERV expression could act as a potential marker of disease severity.

## Conclusions

In this study, we recognised dysregulated expression of endogenous retroviral elements in BALF samples, but not in PBMCs of COVID-19 patients. At the same time, we were able to identify upregulated expression of multiple HERV families in senescence induced HBECs in comparison to non-induced, a fact that could possibly explain the differences in disease severity among age groups. These results indicate that HERV expression might play a pathophysiological role in local inflammatory pathways in lungs afflicted by SARS-CoV-2 and their expression could be a potential therapeutic target.

## Data Availability

The datasets of the PBMCs from COVID-19 patients and healthy volunteers, as well as the data of BALF from COVID-19 patients analysed during the current study are available in National Genomics Data Center-Genome Sequence Archive (NGDC-GSA), in BIG Data Center (https://bigd.big.ac.cn/), Beijing Institute of Genomics (BIG), Chinese Academy of Sciences with accession numbers CRR119890, CRR119891, CRR119892, CRR119893, CRR119894, CRR119895, CRR119896, CRR119897, CRR125445 and CRR125446.

The datasets of BALF from healthy controls analysed in the present work are available in NCBI-Sequence read Archive (SRA), under accession numbers: SRR10571724, SRR10571730, SRR10571732.

The datasets of the induced and non-induced HBECs analysed in the present work are available in NCBI-Sequence read Archive (SRA), under accession numbers: SRR6261633, SRR6261634, SRR6261635, SRR6261636, SRR6261637, SRR6261638, SRR6261639, SRR6261640, SRR6261641, SRR6261642, SRR6261643, SRR6261644, SRR6261645, SRR626163346, SRR6261647, SRR6261648, SRR6261649, SRR6261650, SRR6261651, SRR6261652, SRR6261653, SRR6261654, SRR6261655, SRR6261656.

https://bigd.big.ac.cn/gsa/browse/CRA002390

https://www.ncbi.nlm.nih.gov/sra/?term=srr10571724

https://www.ncbi.nlm.nih.gov/sra/?term=srr10571730

https://www.ncbi.nlm.nih.gov/sra/?term=srr10571732

https://www.ncbi.nlm.nih.gov/sra?term=SRX3367915

https://www.ncbi.nlm.nih.gov/sra?term=SRX3367916

https://www.ncbi.nlm.nih.gov/sra?term=SRX3367917

https://www.ncbi.nlm.nih.gov/sra?term=SRX3367918

https://www.ncbi.nlm.nih.gov/sra?term=SRX3367919

https://www.ncbi.nlm.nih.gov/sra?term=SRX3367920

## Methods

In order to compare the expression of endogenous retroviral elements in the BALF and PBMCs of COVID-19 patients and healthy individuals, online available data were utilized. RNA sequencing (RNAseq) data from BALF samples of 2 COVID-19 patients (2 samples from each) and PBMCs of 3 COVID-19 patients and 3 healthy donors were downloaded in .fastq format from National Genomics Data Center-Genome Sequence Archive (NGDC-GSA) with accession numbers: CRR119890, CRR119891, CRR119892, CRR119893, CRR119894, CRR119895, CRR119896, CRR119897, CRR125445, CRR125446. RNAseq data from BALF samples of healthy individuals were downloaded from Sequence Read Archive (SRA) with accession numbers: SRR10571724, SRR10571730, SRR10571732. All data were paired-end. The platforms used for the production of the raw data were BGISEQ-500 for CRR119890, CRR119891, CRR119892 and CRR119893 (reads of 100 bases length), Illumina MiSeq for the production of CRR119894, CRR119895, CRR119896 and CRR119897 (reads of 150 bases length), Illumina NovaSeq 5000 for the production of CRR125445 and CRR125446 (reads of 150 bases length), and finally Illumina HiSeq 2000 for the production of SRR10571724, SRR10571730 and SRR10571732 (reads of 50 bases length).

For the evaluation of the effect of senescence on the expression of HERV families, data from a non-malignant epithelial oncogene-induced senescence model was used in order to produce senescent HBECs as was described by Komseli et al (11). Non-induced and induced (senescent) HBECs online available RNAseq data were retrieved from NCBI-SRA with accession numbers: SRR6261633, SRR6261634, SRR6261635, SRR6261636, SRR6261637, SRR6261638, SRR6261639, SRR6261640, SRR6261641, SRR6261642, SRR6261643, SRR6261644, SRR6261645, SRR6261646, SRR6261647, SRR6261648, SRR6261649, SRR6261650, SRR6261651, SRR6261652, SRR6261653, SRR6261654, SRR6261655, SRR6261656. For the production of these data Illumina NextSeq500, producing paired-end data (reads of 38 bases length).

### Bioinformatics Analysis

Bowtie2 with default settings for paired-end data was used for the alignment of these data to hg19 human genome assembly (25). Samtools view command with the -q option was used, in order for the mapping quality of the reads included in the alignments to be over 20 (26). Samtools sort and index commands were used with default settings. IGV was used for the visualisation of the mapping alignments (27).

For the identification of the expression of endogenous retroviral elements, the coordinates of HERV-K (HML-2), HERV-H, HERV-W HERV-L, HERV-E, HERV-I, HERV-9, HERV-FRD, HERV-K (HML-1), HERV-K (HML-3), HERV-K (HML-4), HERVK (HML-5) and HERV-K (HML-6) elements were found in the existing literature (28–31). HERV-K (HML-2), HERV-H and HERV-W elements’ coordinates were set in reference genome hg19. Regarding the rest of the endogenous retroviral elements, their coordinates were referring to hg38 assembly and the UCSC Batch Coordinate Conversion (liftOver) online tool was used for the conversion to hg19 coordinates (32).

Bedtools multicov command was used for the quantification of endogenous retroviral expression in each of the datasets used (33). The -f option was used in order to reassure that at least 80% of the read length were overlapping with the HERVs elements’ coordinates. For this reason, based on the coordinates we used and the length of endogenous retroviral elements, we considered a length of 9000bp for HML-3, HML-5 and HML-6, 10000bp for HML-1, HML-2, HML-4 and HML-6, HERV-W, HERV-L, HERV-E, HERV-I and HERV-FRD, a length of 9000bp for HML-3, HML-5 and HML-6, 12000bp for HERV-9 and 17000bp for HERV-H for the calculation of -f for each virus, in order to increase the sensitivity of the detection.

Taking into account the different read lengths and different depths of sequencing among the datasets used in this analysis, we utilised the expression of widely used housekeeping genes for the normalization of our raw reads in each of the datasets. For this purpose, we calculated the expression of SDHA and HPRT1 genes in the datasets used in this analysis. Bedtools multicov command was used for the quantification of the expression of these genes with the -f option for the detection of reads that overlap with the genes coordinates by at least 80%, the same way as with the endogenous retroviral elements. Finally, the sum of the raw read counts from each ERV in each dataset was calculated and was normalized by dividing this number by the expression (in raw reads) of each of the housekeeping genes used, in the respective dataset (normalized expression value). The fold change in the expression of the endogenous retroviral elements in COVID-19 BALF samples and PBMCs was calculated as the ratio of the mean normalized expression value of ERVs in COVID-19 patients to the mean normalized expression value of ERVs in healthy individuals. Respectively, the fold change for the expression between non-induced and induced HBECs was calculated as the ratio of the mean normalized expression value of ERVs in induced (senescent) HBECs to the mean normalized expression value of ERVs in non-induced HBECs.

### Statistical Analysis

IBM Corp. Released 2015. IBM SPSS Statistics for Windows, Version 23.0. Armonk, NY: IBM Corp. was used for the statistical analysis. The normalized expression value of each ERV was log-transformed (ln[normalized expression value]) in order to perform independent sample t-test, with unequal variances assumed (Welch’s t-test), in order to identify statistically significant differences between the BALF samples and PBMCs of COVID-19 patients and healthy controls as well as to identify statistically significant differences in the expression of the studied HERVs between HBECs without and with oncogene-induced senescence.

**List of Abbreviations**
COVID-19: (Corona Virus Disease 2019)
SARS-CoV: (Severe Acute Respiratory Syndrome Coronavirus)
HERVs: (Human Endogenous Retroviruses)
PBMCs: (Peripheral Blood Monocytes)
BALF: (Bronchoalveolar Lavage Fluid)
HBECs: (Human Bronchial Epithelial Cells)
SARS-CoV-2: (Severe Acute Respiratory Syndrome Coronavirus 2)
SDHA: (Succinate Dehydrogenase) Hypoxanthine Phosphoribosyl Transferase 1 (HPRT1)
HERV-W: *env* (HERV-W envelope protein)
hg19: (human genome, version 19)
hg38: (human genome, version 38)
ERV: (Endogenous retrovirus)

## Declarations

### Ethics Approval and Consent to Participate

Not applicable

### Consent for Publication

Not applicable

### Availability of Data and Materials

The datasets of the PBMCs from COVID-19 patients and healthy volunteers, as well as the data of BALF from COVID-19 patients analysed during the current study are available in National Genomics Data Center-Genome Sequence Archive (NGDC-GSA), in BIG Data Center (https://bigd.big.ac.cn/), Beijing Institute of Genomics (BIG), Chinese Academy of Sciences with accession numbers CRR119890, CRR119891, CRR119892, CRR119893, CRR119894, CRR119895, CRR119896, CRR119897, CRR125445 and CRR125446 (https://bigd.big.ac.cn/gsa/browse/CRA002390) (9).

The datasets of BALF from healthy controls analysed in the present work are available in NCBI-Sequence read Archive (SRA), under accession numbers: SRR10571724 (https://www.ncbi.nlm.nih.gov/sra/?term=srr10571724), SRR10571730 (https://www.ncbi.nlm.nih.gov/sra/?term=srr10571730), SRR10571732 (https://www.ncbi.nlm.nih.gov/sra/?term=srr10571732).

The datasets of the induced and non-induced HBECs analysed in the present work are available in NCBI-Sequence read Archive (SRA), under accession numbers: SRR6261633, SRR6261634, SRR6261635, SRR6261636 (https://www.ncbi.nlm.nih.gov/sra?term=SRX3367915), SRR6261637, SRR6261638, SRR6261639, SRR6261640 (https://www.ncbi.nlm.nih.gov/sra?term=SRX3367916), SRR6261641, SRR6261642, SRR6261643, SRR6261644 (https://www.ncbi.nlm.nih.gov/sra?term=SRX3367917), SRR6261645, SRR626163346, SRR6261647, SRR6261648 https://www.ncbi.nlm.nih.gov/sra?term=SRX3367918), SRR6261649, SRR6261650, SRR6261651, SRR6261652 (https://www.ncbi.nlm.nih.gov/sra?term=SRX3367919), SRR6261653, SRR6261654, SRR6261655, SRR6261656 (https://www.ncbi.nlm.nih.gov/sra?term=SRX3367920).

## Acknowledgements

The study is supported by:

GSRT No 2020ΣE01300001

Emblematic action to handle SARS-CoV-2 infection: Epidemiological study in Greece via extensive testing for viral and antibody detection, sequencing of the virome and genetic analysis of the carriers (GSRT No 2020ΣE01300001).

---

<u>Additional File 1:</u> File Format: Tagged Image File Format (.tif)

Title of Data: Figure 5: No statistically significant differences were observed in the expression of HERV-K (HML-1, HML-2, HML-3, HML-4 HML-5 and HML-6) between healthy donors and COVID-19 patients (SDHA normalization).

Description of Data: Expression of HERV-K (HML-1), HERV-K (HML-2), HERV-K (HML-3), HERV-K (HML-4), HERV-K (HML-5), HERV-K (HML-6) in PBMCs of healthy controls and COVID-19 patients normalized with the expression of SDHA gene. No statistically significant differences were observed between healthy donors and COVID-19 patients.

<u>Additional File 2:</u> File Format: Tagged Image File Format (.tif)

Title of Data: Figure 6: Downregulation of the expression of ERV-L and HERV-FRD in the PBMCs of COVID-19 patients (SDHA normalization).

Description of Data: Expression of HERV-W, HERV-H, HERV-L, HERV-9, HERV-E, HERV-FRD and HERV-I in PBMCs of healthy controls and COVID-19 patients normalized with the expression of SDHA gene. Statistically significant downregulation of the expression of ERV-L (0.85 fold change, *p=0.029*) and HERV-FRD (0.64 fold change, *p=0.049*) was observed in the PBMCs of COVID-19 patients in comparison to PBMCs of healthy donors.

<u>Additional File 3:</u> File Format: Tagged Image File Format (.tif)

Title of Data: Figure 7: Downregulation in the expression of HML-1 in the PBMCs of COVID-19 patients (HPRT1 normalization).

Description of Data: Expression of HERV-K (HML-1), HERV-K (HML-2), HERV-K (HML-3), HERV-K (HML-4), HERV-K (HML-5), HERV-K (HML-6) in PBMCs of healthy controls and COVID-19 patients normalized with the expression of HPRT1 gene. Statistically significant downregulation was observed in the expression of HML-1 (0.74 fold change, *p=0.007*) in the PBMCs of COVID-19 patients in comparison to PBMCs of healthy donors.

<u>Additional File 4:</u> File Format: Tagged Image File Format (.tif)

Title of Data: Figure 7: Figure 8: No statistically significant differences were observed in the expression of HERVs between healthy donors and COVID-19 patients.

Description of Data: Expression of HERV-W, HERV-H, HERV-L, HERV-9, HERV-E, HERV-FRD and HERV-I in PBMCs of healthy controls and COVID19 patients normalized with the expression of HPRT1 gene. No statistically significant differences were observed between healthy donors and COVID-19 patients.

